# Impact of Systemic Arterial Pressure, Collateral Vascular Resistance and Degree of Carotid Stenosis on Cerebral Blood Flow, Reserve Blood Flow, Critical Carotid Stenosis, Cerebral Ischemia and Carotid Hemodynamics

**DOI:** 10.1101/2021.02.23.21252300

**Authors:** Joseph P Archie

## Abstract

**Introduction:** Carotid artery stenosis related stroke is a major health care concern. Current risk management strategies for patients with asymptomatic carotid stenosis include ultrasound surveillance and occasionally an estimate of cerebral blood flow reserve. Other patient specific hemodynamic variables may be predictive of ischemic stroke risk. This study, based on a cerebral blood flow hemodynamic model, aims to investigate the impact of systemic arterial pressure, collateral vascular resistance and degree of carotid stenosis on cerebral ischemic risk, cerebrovascular blood flow reserve, critical carotid artery stenosis, carotid artery blood flow and carotid stenosis hemodynamics.

**Methods:** This study uses a three-component (carotid, collateral, brain) energy conservation cerebrovascular fluid mechanics model in combination with the Lassen cerebral blood flow autoregulation model that predicts cerebral blood flow in patients with carotid stenosis. It is a two-phase model, zone A when regional cerebral blood flow is autoregulated at normal values and zone B when cerebral blood flow is below normal and dependent on collateral perfusion pressure. The model solution with carotid artery occlusion defines collateral vascular resistance, with patient specific values calculated from clinical pressure measurements. In addition to cerebral blood flow the model predicts critical stenosis values and carotid and collateral blood flows as a function of systemic arterial pressure and percent diameter stenosis. Carotid stenosis blood flow velocities and energy dissipation are predicted from carotid blood flow solutions.

**Results:** The model defines patient specific collateral vascular resistance, cerebral vascular resistance and critical carotid stenosis. It predicts carotid vascular resistance to be non-linearly proportional to area carotid stenosis. Solutions include reserve cerebral blood flow, the carotid and collateral components of cerebral blood flow, criteria for cerebral ischemia and carotid stenosis hemodynamics. Critical carotid stenosis is determined by mean systemic arterial pressure and the Lassen autoregulation threshold cerebral perfusion pressure. Critical stenosis values range from 61% to 76% diameter stenosis when mean systemic arterial pressures are 80mmHg to 120mmHg and the cerebral autoregulation pressure threshold is 50mmHg. When carotid stenosis is less than critical, cerebral blood flow is maintained normal and the ratios of carotid blood flow to collateral blood flow are inversely proportional to the carotid to collateral vascular resistance ratios. At stenosis greater than the critical, carotid blood flow is not adequate to maintain normal cerebral blood flow, cerebral blood flow is primarily collateral flow, all reserve blood flow is collateral and prevention of cerebral ischemia requires adequate collateral flow. Patient specific collateral vascular resistance values less than 1.0 predict normal cerebral blood flow at moderate to severe stenosis. Values greater than 1.0 predicts cerebral ischemia to be dependent on the magnitude of collateral vascular resistance. Systemic arterial pressure is a major determinant of carotid stenosis hemodynamics. Carotid blood flow velocities increase with carotid stenosis and have progressively higher variance depending on collateral blood flow as predicted by collateral vascular resistance. Turbulent flow energy dissipation intensity is similarly inversely proportional to collateral vascular resistance at severe carotid stenosis.

**Conclusions:** Cerebral, collateral and carotid blood flow solutions are determined by systemic arterial pressure, collateral vascular resistance and degree of stenosis. Critical carotid stenosis, systemic arterial pressure and collateral vascular resistance are primary determinants of cerebral ischemic risk in patients with significant carotid stenosis.

## Introduction

Carotid artery stenosis related stroke is a major health care concern. Current risk management strategies for patients with asymptomatic carotid stenosis include ultrasound surveillance and occasionally an estimate of cerebral blood flow reserve. Other patient specific hemodynamic variables may be predictive of ischemic stroke risk.

The term “critical carotid stenosis” is often used but rarely defined. To clinicians “critical” usually implies a degree of carotid artery narrowing associated with an increased risk of stroke, usually considered to be severe 70% to 99% diameter stenosis. Critical carotid stenosis means the maximum degree of carotid stenosis at which adequate or normal cerebral blood flow can be maintained. However, clinical and experimental studies indicate that normal cerebral blood flow is maintained by the collateral circulation in many patients with severe carotid stenosis or occlusion. The most precise definition of critical carotid stenosis is the degree of stenosis beyond which internal carotid blood flow alone fails to maintain normal cerebral blood flow. Many authors correctly argue that critical arterial stenosis depends on depends on several independent variables including blood flow. While this is true for arterial stenosis in general, the three-component carotid-collateral-cerebral vascular system model when combined with the Lassen autoregulation model used here defines and predicts patient specific values of critical carotid stenosis. The key to predicting cerebral ischemic risk in patients with asymptomatic carotid stenosis is their critical stenosis value. The aims of this study using a hemodynamic model are to investigate the impact of systemic arterial pressure, collateral vascular resistance and carotid stenosis on cerebral ischemia, cerebrovascular reserve, critical carotid stenosis, carotid artery blood flow and carotid stenosis hemodynamics.

## Methods

This study combines clinical results with a basic one-dimensional hemodynamic energy conservation model of the cerebrovascular system in man. Physics and engineering laws and principals are equally applicable to biological systems. The three conservation laws are mass, momentum and energy. Each of these has been used to describe hemodynamic systems in man that include arterial stenosis. Constitutive rules or equations are necessary to connect the forces, pressures or stresses to the motion, displacements or strains in the conservation of momentum and conservation of energy law. Many constitutive equations are simple and linear. For example, when the law of conservation of momentum is applied to a solid material the ratio of force or stress to displacement or strain is a material specific elastic-viscoelastic material-constitutive property, like Young’s modulus. For a simple fluid, the constitutive connection between pressure and flow is viscosity. For example, a Newtonian fluid has a fixed/constant viscosity. A similar constitutive coupling term or equation is necessary for the law of conservation of energy. The energy model developed by Bernoulli neglected viscosity and is the basis of hydrodynamics or hydrology. When the law of conservation of energy is applied to a hemodynamic system the vascular resistances are the constitutive values/functions that couple the pressure gradients to the blood flow. The cerebrovascular energy model given in Figure 1a has three constitutive resistances; carotid, brain and collateral (Rc, Rb and Rw). The models given in Figures 1 a, b.c when combined with the Lassen cerebral blood flow autoregulation model in Figure 2, predict patient specific carotid blood flows and carotid stenosis Reynolds numbers, velocities and energy dissipation as a function percent diameter carotid stenosis. The model also defines the three constitutive vascular resistance values/functions. This hemodynamic model has been used to predict cerebral blood flow and reserve blood flow (1, 2).

**Figure 1a,b,c.**
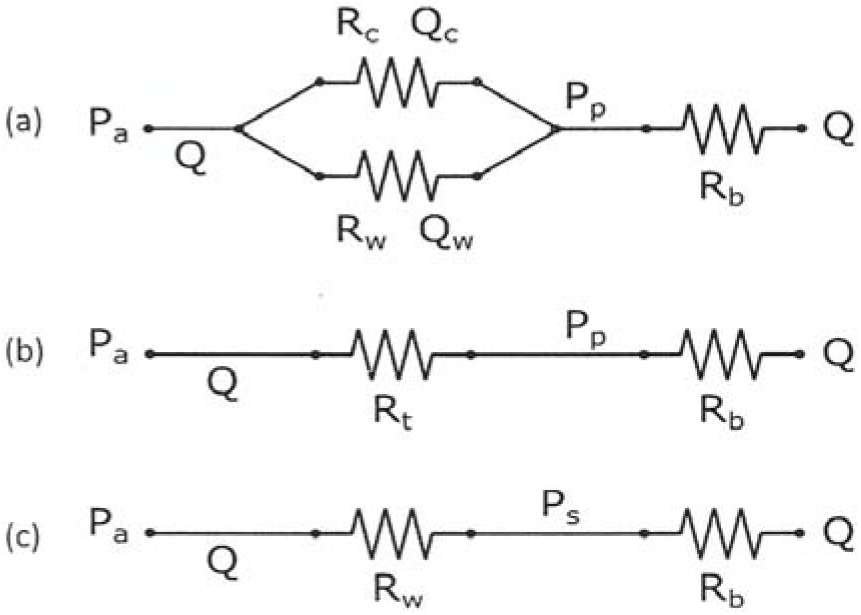
Vascular energy model schematics. (a) The three-component energy model for cerebral blood flow. Pa is mean systemic arterial pressure, Rc is carotid stenosis vascular resistance and Rw is collateral vascular resistance. Qc and Qw are the parallel carotid and collateral blood flows with cerebral blood flow Q = Qc + Qw. Pp is cerebral perfusion pressure. Rb is brain vascular resistance. When Pp is greater than the Lassen lower threshold, pressure (Figure 2) Q is normal autoregulated cerebral blood flow, Qn. When Pp is less than the threshold Q is less than Qn, cerebral ischemia. The model has two components; when Qn is autoregulated normal in Zone A and when Q is pressure dependent in zone B. (b) The model with the parallel resistors combined, Rt = RcRw/(Rc + Rw). (c)The model when carotid stenosis is 100% (Rc = ∞, occlusion). The cerebral perfusion pressure, Pp, is carotid stump pressure, Ps, measured clinically. A model solution defines patient specific collateral vascular resistance, Rw.

**Figure 2.**
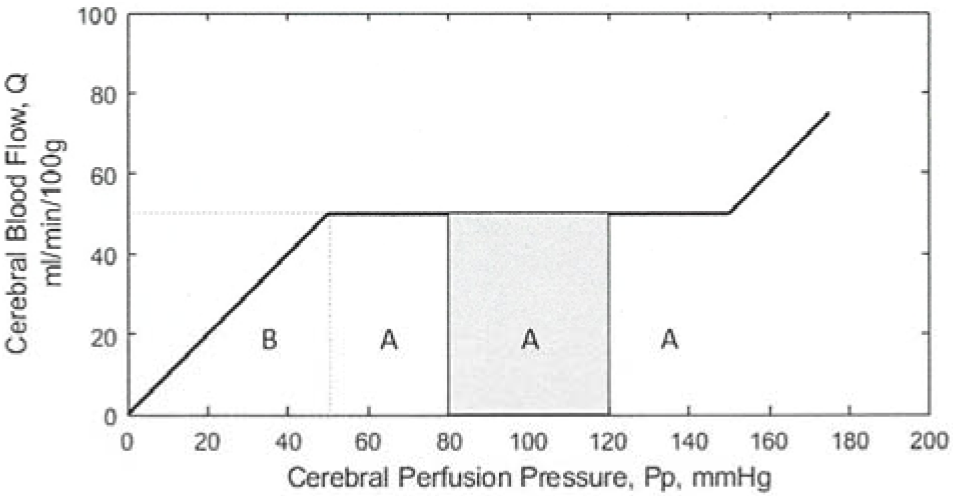
Schematic of the Lassen cerebral blood flow autoregulation model (3, 4). When combined with in the energy hemodynamic model in Figure 1, cerebral blood flow in zone A is autoregulated normal and in zone B cerebral blood flow is pressure dependent and below normal.

## Cerebral Vascular Resistance

This is defined by combining the energy conservation model with the cerebral autoregulation model. The generally accepted brain blood flow autoregulation model was originally published by Lassen (3, 4) as given in Figure 2. In his 1959 description of cerebral auto-regulation in man (3) the range of mean cerebral perfusion pressures over which blood flow was determined to be maintained at a constant normal rate has a lower threshold of 50 mmHg and extends to approximately 150 mmHg. He also determined normal regional cerebral blood flow over this perfusion pressure range to be 50 millimeters per minute per 100 grams of brain, or 50/ml/min/100g. This value has been confirmed (5), as has a threshold regional cerebral blood flow values of 30ml/min/100g threshold for cerebral symptoms and 18ml/min/100g threshold for cerebral infarction (6, 7). In a later publication a lower threshold cerebral perfusion pressure to 60 mmHg was suggested to include hypertensive patients (4). However, many investigators continue to use 50mmHg. Incorporating the Lassen autoregulation model into the hemodynamic model results in two interfaced cerebral blood flow zones, normal autoregulation blood flow designated as zone A and below normal cerebral blood flow designated zone B. The lower threshold pressure for autoregulation determines the interface, here in assumed to be 50mmHg. The Lassen model cerebral vascular resistance, Rb, It is a variable in zone A and a constant in zone B.

### Collateral Vascular Resistance

This is defined by a model solution to the energy conservation model when the carotid is occluded, Figure 1b. Several decades ago it was recognized that carotid artery stump pressures measured during surgery with proximal carotid clamping was an estimate of collateral flow potential. It was also found that pharmacologic induction of mild systemic hypertension increased carotid stump pressure. This was occasionally used to elevate very low carotid stump pressures to avoid use of a shunt during carotid surgery. Archie and Feldtman (8) initially considered collateral vascular resistance to be the ratio of mean systemic pressure to carotid stump pressure, Pa/Ps. They later found this ratio to be patient specific over a range of perfusion pressures (9). An exact definition of collateral vascular resistance (Rw) is obtained from the model solution when the carotid is occluded and cerebral perfusion pressure is carotid stump pressure (Pp = Ps) as given in Figure 1c. The equation is Rw = Rb(Pa/Ps – 1). In the pressure dependent zone B, Rb = 1.0 and Rw = (Pa/Ps – 1). In zone A, Rb = Ps/Qn, and collateral vascular resistance is Rb = Ps/Qn and Rw = Ps/Qn(Pa/Ps – 1) or Rw = (Pa – Ps)/Qn. In this study Qn = 50ml/min/100g, and Rw = (Pa – Ps)/50.

### Carotid Vascular Resistance

As given above, the combined energy and Lassen models define the two constitutive functions, brain and collateral vascular resistance, Rb and Rw. Similarly, the third constitutive function, carotid vascular resistance, Rc, is defined by the model. The three resistances must be internally consistent with the model to satisfy the laws of fluid mechanics and clinical observations in patients with carotid stenosis. Carotid vascular resistance as a function of carotid stenosis must satisfy the end-point values of zero in the absence of carotid stenosis (0 % stenosis) and infinity at carotid occlusion (100 % stenosis). The model equation is Rc = (Pa – Pp)/Qc, (Figure 1), where Pa is mean systemic arterial pressure, Pp is cerebral perfusion pressure and Qc is the carotid blood flow contribution to cerebral blood flow, Q. Experimental investigations of arterial stenosis determined that volume flow rates decreased in proportion to percent area stenosis (10). That is, that the magnitude of blood flow, Qc, is inversely proportional to percent area stenosis. That means that Qc = 1/(% area stenosis) = 1/(1 – A2/A1), where A1 is normal vessel cross-sectional area and A2 is stenosis area (assuming a round stenosis lumen). It was also found that decreasing stenosis lumen area produces a non-linear increase in the pressure gradient across the stenosis (11). Thus, (Pa – Pp) = 1/(A2/A1), and Rc = (Pa – Pp)/Qc = (1- A2/A1)/(A2/A1). The term (1- A2/A1) is fractional percent area stenosis and is equal to [1 – (D2/D1)^2^], fractional percent diameter stenosis where A = 4 D^2^/_π_ and (A2/A1) = (D2/D1)^2^. In this study the model results are given as a function of percent diameter stenosis, X, where X = (D2/D1) and X^2^= (A2/A1). Therefore Rc = X^2^/(1 + X^2^) or Rc = 1/[(1/X^2^) - 1], where X is fractional diameter percent carotid stenosis. The experimental results strongly support a non-linear relationship of power two. This model predicted function for carotid vascular resistance satisfies experimental and clinical observations.

### The Complete Model

Combining the three component of the one-dimensional energy model (Figure 1a) with the Lassen autoregulation model (Figure 2) gives a set of algebraic equations for cerebral blood flow. In addition to the cerebral blood flow solutions the model defines the relationships between three patient specific independent variables; systemic arterial pressure, degree of diameter carotid stenosis and collateral vascular resistance (1, 2). A typical solution is given in Figure 3 when collateral vascular resistance values (Rw) are ∞, 3.0, 1.0 and 0.5 and systemic arterial pressure is 100mmHg. The upper “reserve flow” component is the cerebral blood flow autoregulated model zone

**Figure 3.**
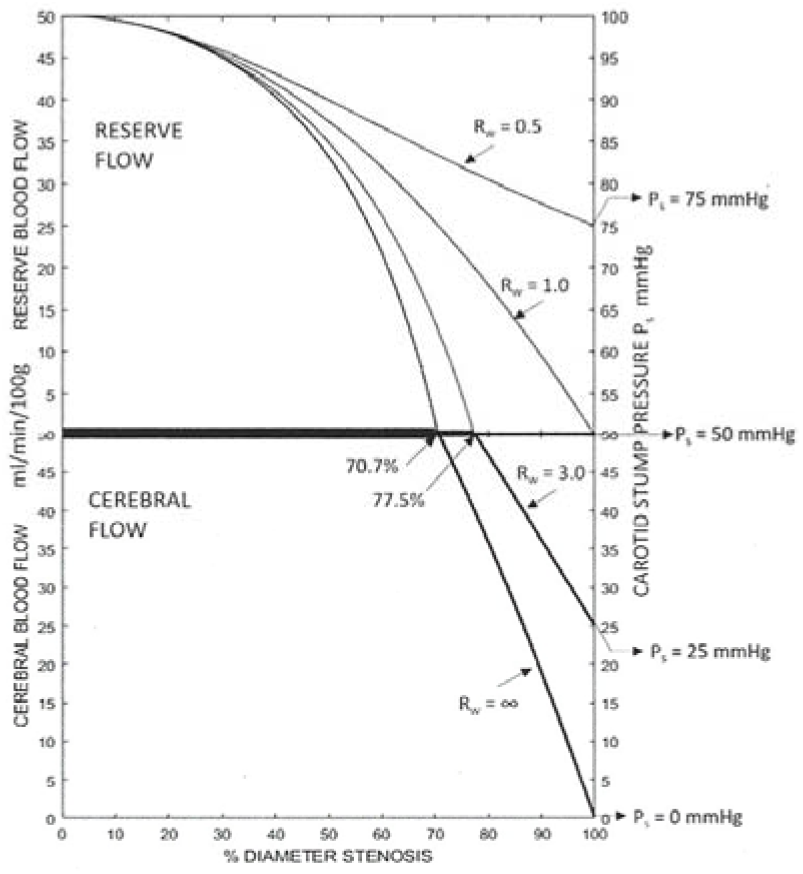
Cerebral blood flow solutions for carotid blood flow alone (Rw = ∞)and three patient specific collateral vascular resistances. Systemic arterial pressure is 100mmHg. In the theoretical absence of collateral blood flow cerebral ischemia at 70 to 75% stenosis. The mean patient collateral vascular resistance is Rw = 1.0. This is adequate collateral flow to prevent cerebral ischemia at carotid occlusion but no remaining reserve. Patients at the lower standard deviation Rw = 0.5 have excellent collateral flow with reserve at carotid occlusion. Conversely, patients at the upper standard deviation Rw = 3.0 have little collateral flow and develops cerebral ischemia above 77% stenosis.

A. The lower “cerebral flow” component is the pressure dependent cerebral blood flow model zone B. The interface is normal cerebral blood flow of 50ml/min/100g until carotid diameter stenosis Rc value combined with Rw makes Rt = 1.0 (Figure 1b, Rt = RcRw/[Rc + Rw], and Pa = [Rt + 1]Qn), the end of autoregulation. For example when Rw = ∞, it is 70.7% stenosis, when Pa = 100mmHg. For Rw = 3.0 it is 77.5% and for Rw = 1.0 it is 100%. For Rw = 0.5 there is reserve cerebral blood flow of 25ml/min/100g at carotid occlusion. This explains why in general the model predicts adequate cerebral blood flow in the approximate 50% of patients with Rw < 1.0 that progress to carotid occlusion. It is the other about 50% of patients with Rw > 1.0 that this study is concerned with.

## RESULTS

### Cerebral Vascular Resistance

Many human organs and regions like the extremities regulate blood flow by adjusting local vascular resistance to meet current oxygen demand. The normal brain is unique in that it autoregulates cerebral blood at normal value of approximately 50ml per minute per 100 grams of brain over a wide range of cerebral perfusion pressures. The Lassen model in Figure 2 illustrates this where the lower threshold of autoregulation perfusion pressure is 50mmHg. At normal human mean systemic pressure of 90mmHg to 110mmHg, a 40mmHg to 50mmHg reduction in cerebral perfusion pressure is necessary to reach the lower Lassen 50mmHg threshold. When cerebral perfusion pressure is greater than the 50mmHg (zone A) threshold cerebral blood flow is autoregulated normal and there is reserve blood flow. Below this perfusion pressure threshold cerebral blood flow is impaired and linearly dependent on cerebral perfusion pressure (zone B). Cerebral vascular resistance is the ratio of perfusion pressure to blood flow, or Rb = Pp/Q. At the Lassen model threshold, Pp = 50mmHg, Qn = 50 ml/min/100g and Rb = 50/50 = 1.0 in zone B. When cerebral perfusion is greater than the 50mmHg Lassen model mean systemic arterial pressure, Rb = Pp/50. For example, if Pa = 90mmHg, Rb = 90/50 = 1.8. The interface between the autoregulation model, zone A, and the pressure dependent model, zone B, is the Lassen threshold 50mmHg.

Carotid stenosis produces a pressure gradient making brain vascular resistance, Rb, a variable from Pa/50 to Pp/50, depending on percent stenosis. If Pp reaches the 50mmHg threshold Rb = 1.0. For a Lassen autoregulation cut-point of 60mmHg, Rb = Pa/60 at zero stenosis and at minimum Rb = 1.2.

### Collateral Vascular Resistance

Several decades ago it was recognized that carotid artery stump pressures measured during carotid clamping was an estimate of patient specific collateral blood flow potential. It was also noted that mild systemic hypertension increases carotid stump pressure. This was occasionally used to elevate very low carotid stump pressures to theoretically avoid use of a shunt during the carotid surgery. It was further recognized that the systemic arterial pressure to carotid stump pressure ratio, Pa/Ps, was an index of collateral vascular resistance (8) and was patient specific (9). An accurate solution for collateral vascular resistance was obtained by solving the model equation for Figure 1c. When there is no carotid artery blood flow carotid vascular resistance Rc = ∞ and fractional percent diameter carotid stenosis X = 1.0. Cerebral perfusion pressure Pp is the carotid stump pressure Ps, and the solution is Rw = Rb(Pa/Pp – 1). When Ps is less than the 50mmHg threshold autoregulation, zone B, Rb = 1 and Rw = (Pa/Ps - 1). When carotid stump pressure is greater than the autoregulation threshold, zone A, Rb = Ps/Qn and Rw = (Ps/Qn)(Pa/Ps - 1) or Rw = (Pa – Ps)/Qn. As Qn = 50ml/min/100gg. Rw = Ps/50(Pa/Ps – 1) or Rw = (pa – Ps)/50. Mean and +/-1SD of Rw values were determined from 1, 260 clinically measured Pa and Ps values (12). The approximate values are mean Rw = 1.0, upper one standard deviation Rw = 3.0 and lower one standard deviation Rw = 0.5. The Rw values 0.5 to 3.0 include approximately two thirds of the patients in the population having carotid surgery for stenosis. The three numerical Rw values are used to illustrate the between patient variance of results.

### Carotid Vascular Resistance

If the combined energy conservation model (Figure 1) and Lassen auto-regulation model (Figure 2) has a potentially weak link it is carotid vascular resistance, Rc. The model generated function for carotid vascular resistance is Rc = 1/[(1/X^2^) - 1] where X is fractional percent diameter stenosis. The equation for Rc must be internally consistent in the model with Rb and Rw. For example, when Pa = 100mmHg, Rb normally decreases from 2.5 at zero stenosis to a minimum of 1.0 if cerebral perfusion is reduced to the Lassen 50mmHg threshold at the intersection of zones A and B. The Rc equation has an X range of 0 to 1.0, (0% to 100% diameter stenosis). If Rb decreases to 1.0 all carotid blood flow reserve is depleted and further stenosis reduces the carotid contribution to cerebral blood flow below normal. For patients with collateral vascular resistance Rw > 1.0, the model equation is Pa = (Rt + Rb)Qn and assuming Pa = 100mmHg, Rt = [(100/50) – 1] = 1.0. For patients with a carotid stump pressure of 25mmHg, Rc = 1/[(1/Rt – 1/Rw), Rc = 1.50, and X = 0.775 or 77.5% stenosis diameter stenosis. Similarly, when Ps =18mmHg, the cerebral blood flow infarction threshold, Rc = 1.78, X = 0.75 or 75%. Both of these values are internally consistent with the Rc model.

Most published studies of carotid stenosis vascular resistance fail to consider turbulence or that it can develop at mild to moderate carotid stenosis. While the standard fluid mechanics Navier-Stokes equation are applicable to turbulent flow the only valid solutions are limited to laminar flow. Some theoretical studies of arterial stenosis use the perfect fluid Bernoulli kinetic energy or the laminar flow Poiseuille solutions. Unfortunately both give a fourth power function of percent diameter stenosis. While this may be accurate at small pressure gradients due to fluid viscosity, the large energy loss with turbulence at high pressure gradients and low brain perfusion pressures due moderate to severe carotid stenosis are not taken into account.

Two generally accepted clinical observations are the onset of a cervical bruit as well as onset of turbulent flow near 50% diameter stenosis and the increased risk of cerebral ischemia near the 70% diameter stenosis transition from “moderate” to severe stenosis.

Cerebral ischemia occurs when cerebral pressure gradient is less the Lassen lower 50mmHg autoregulation threshold. For example, at systemic arterial pressure of 100mmHg, Rc = (100-50)/5 = 1.0 and X = 0.71 or 71% diameter stenosis. Using the fourth power of X in the Rc gives 80% and 90% stenosis, far above clinical results. The stenosis area Rc model is further supported by the results.

### Critical Carotid Stenosis

Critical carotid artery stenosis is defined as the percent diameter stenosis at which cerebral blood flow ceases to be auto-regulated and becomes pressure dependent in the absence of collateral blood flow. The Lassen lower threshold cerebral perfusion pressure determines cerebral perfusion pressure at this junction. The solution to the model equation in Figure 1c when Q = Qn gives the critical carotid stenosis values. The solution is Pa = (Rc + Rb)Qn, or Rc = Pa/Qn - Rb. Assuming the Lassen threshold is 50mmHg, Rc = Pa/50-1.0. For Pa values of 80, 90, 100, 110, 120 mmHg the Rc values are 0.6, 0.8, 1.0, 1.2, and 1.4 respectively giving critical percent diameter stenosis values (100X) of 61.2%, 66.7%, 70.7%, 73.9% and 76.4% respectively. If a Lassen threshold of 60mmHg is used, the critical percent stenosis values range from 53.7% to 73.9% for the same mean arterial pressure values. Archie and Feldtman (13) measured internal carotid artery blood flow intra-operatively before and after carotid endarterectomy in 39 patients to determine the critical carotid stenosis in this cohort as given in Figure 4. The critical carotid stenosis value based on the regression line was 59.6% diameter. The patient’s mean systemic arterial pressures varied during surgery had an average value near 90mmHg. The model predictions of critical percent diameter stenosis for Pa = 90mmHg is 66.7%; close to the clinical measured 59.6%.

**Figure 4.**
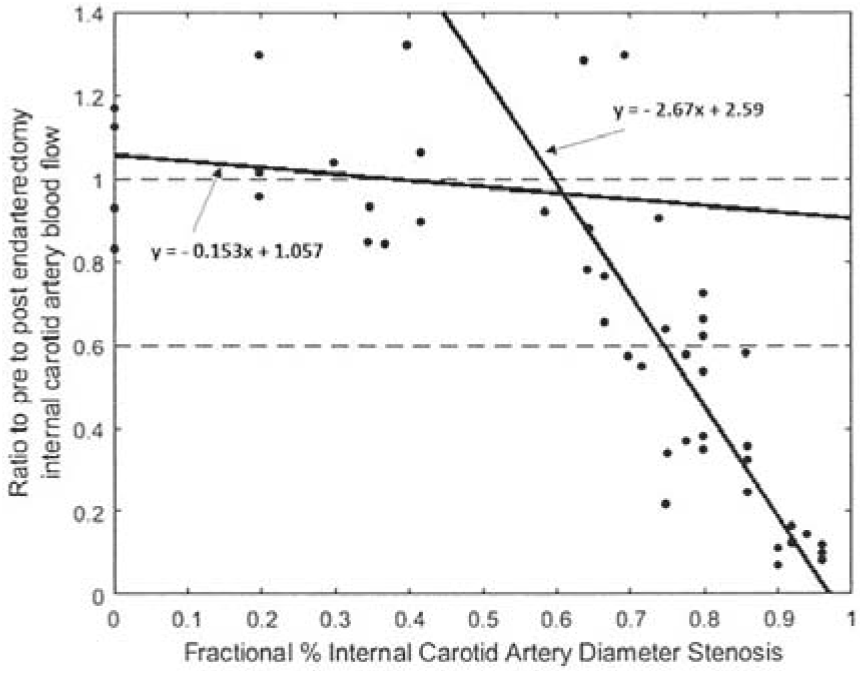
Critical carotid stenosis determined from measurements of internal carotid blood flow and percent diameter stenosis in 39 patients (10). The critical stenosis intercept is approximately 60%.

The 61% to 76% diameter range of critical carotid stenosis above which normal cerebral blood flow is not maintained by the carotid artery alone is consistent with clinical measurements and estimates. Collateral blood flow becomes the dominant contributor to cerebral blood flow in patients with carotid stenosis greater than the critical value. A review and Meta analysis of patients with greater than 70% diameter stenosis found a significantly higher probability of ischemic stroke or TIA in 3-year follow-up if they do not have some measurable cerebral blood flow reserve or reactivity (14). The authors conclude that maintenance of normal cerebral blood flow, with or without reserve blood flow, is due entirely to the availability of collateral flow. Similarly, reserve blood flow predictions based on this hemodynamic model are consistent with these findings (2). Clinical evidence supports the concept that carotid critical stenosis is in the 50% to 75% range and most often near 60% to 65%, depending on systemic arterial pressure.

### Systemic Arterial Pressure

Systemic arterial pressure is a primary determinant of patient specific collateral vascular resistance, Rw = Rb(Pa/Ps −1). When Rw is greater than 1.0 cerebral perfusion pressure is reduced to the Lassen threshold of 50mmHg at carotid stenosis equal to or greater than greater the critical value. This is illustrated in Figure 5 for mean systemic pressures from 80mmHg to 120mmHg. The Rw values are ∞, 5, 2 and 1.

**Figure 5.**
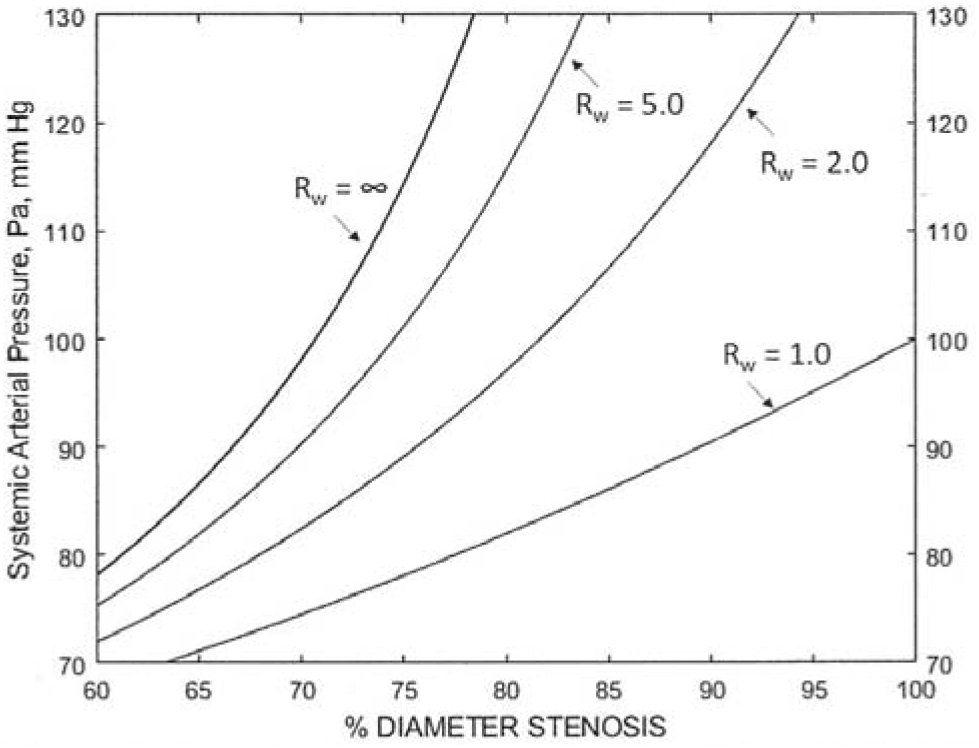
The effect of systemic arterial pressure, Pa, and collateral vascular resistance, Rw, on percent carotid stenosis at which reserve cerebral blood flow is depleted and cerebral flow remains normal, Q = 50mmHg. This is the model interface between the autoregulation A zone and the pressure dependent B zone. Systemic arterial pressure, Pa, ranges from 80mmHg to 120mmHg and collateral vascular resistance, Rw, values are 5.0, 2.0 and 1.0. The Rw = ∞ curve is when there is no collateral blood flow. The approximate mean patients value is Rw = 1.0. Patients with poor collateral blood flow have Rw > 1.0.

When cerebral perfusion pressure is less than 50mmHg and Rw is greater than 1.0, the influence of systemic pressures of 80mmHg to 120mmHg on impaired cerebral blood flow is illustrated in figure 6. The large dots on the Q = 50 threshold horizontal line are the critical stenosis values for 80mmHg and 12mmHg. In the lower frame collateral vascular resistance is 5.0 and increasing systemic pressure from 80 to 120mmHg raises cerebral blood flow from 12.5 ml/min/100g to 20 ml/min/100g, slightly above the infarction threshold of 18mm/min/100g. In the upper frame Rw = 3.0 and at carotid occlusion cerebral blood flow increases from 20 to 30ml/min/100g, the threshold for onset of symptoms. Higher mean systemic arterial pressures decrease the percentage of patients with impaired cerebral blood flow at high-grade stenosis or occlusion. Similarly, increasing a patients systemic pressure is predicted to decrease their probability of cerebral ischemia if they have poor collateral circulation. Perhaps mild hypertension is protective in these patients while systemic hypotension is not well tolerated even in some patients with collateral vascular resistance values less than 1.0.

**Figure 6a,b.**
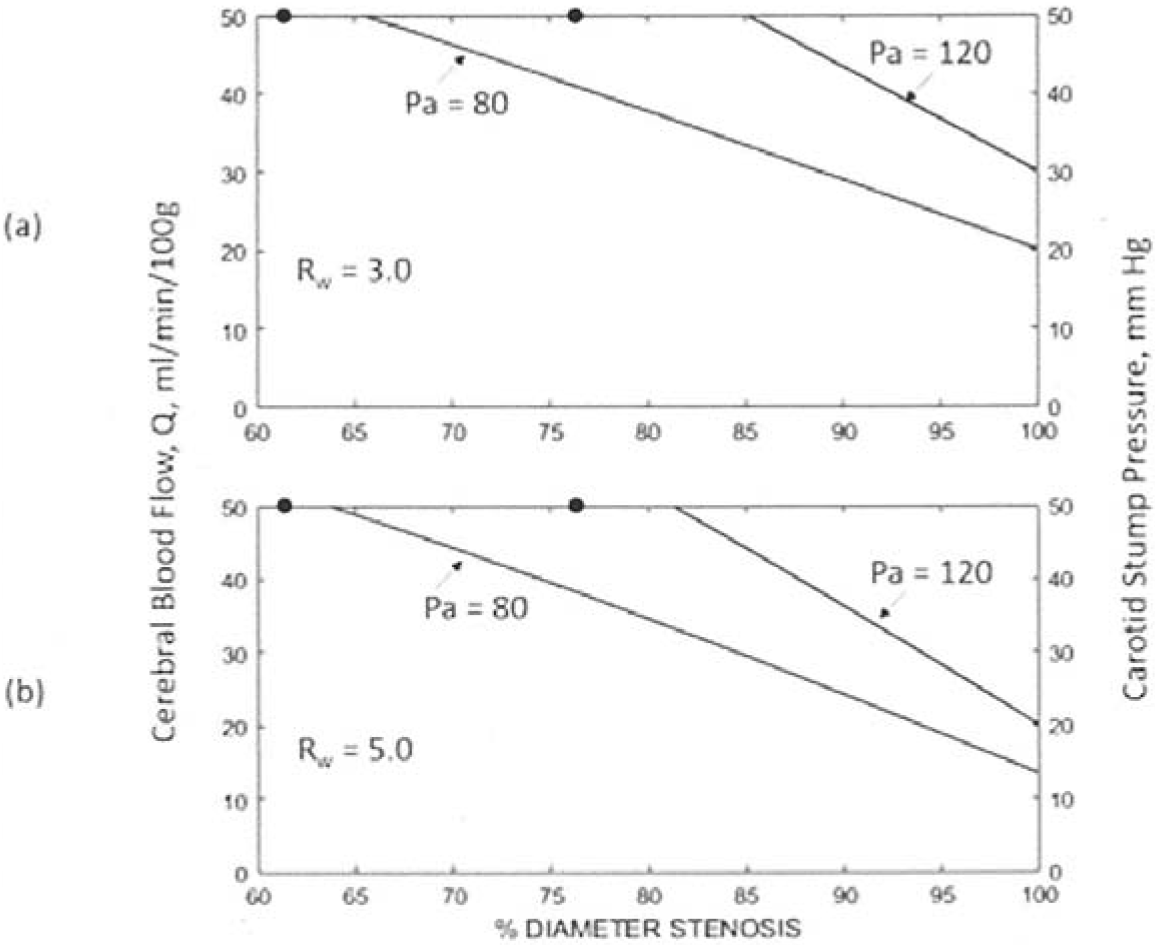
The effect of systemic arterial pressures 80mmHg to 120mmHg on the degree of impaired cerebral blood flow as a function of percent diameter stenosis in patients with poor collateral vascular resistance. The dots on the Q = 50 interface between the A and B model zones are critical carotid stenosis values for Pa = 80 and Pa = 120mmHg. (a) Collateral vascular resistance Rw = 3.0. Approximately 16% of patients will have a Rw value > 3.0 and impaired cerebral blood flow in the symptomatic ischemic zone below 30ml/min/100g. (b) Collateral vascular resistance Rw =5.0. Approximately 9% of patients will have Rw values > 5.0 and many will have cerebral blood flow less than the 18 ml/min/100g infarction threshold at carotid occlusion depending on systemic arterial pressure, Pa.

### Carotid Blood Flow

The risk of cerebral ischemic symptoms or stroke in patients with carotid artery stenosis is predicted to be dependent on the combined effects of critical carotid stenosis, collateral vascular resistance and carotid blood flow.Determining the carotid, Qc, and collateral, Qw, blood flow contributions to cerebral blood flow Q (Q = Qc + Qw) is complicated. The carotid and collateral blood flow systems are anatomically parallel in humans and as in the model. Carotid blood flow reserve expires at critical carotid stenosis and with further increase in carotid stenosis the carotid contribution to cerebral blood flow is not sufficient to prevent cerebral ischemia. Carotid blood flow becomes linearly dependent on cerebral perfusion pressure at higher degrees of stenosis (zone B). This is illustrated in Figure 7a when Pa = 100mmHg and all cerebral blood flow is from the carotid. The Rw = ∞ result is theoretical because to date no patient has been found to have an accurately measured carotid stump pressure of zero. However, it serves as a guideline for understanding the results for patient’s Rw values. When collateral blood flow is a finite value there are three options. Collateral vascular resistance, Rw, is either inadequate to prevent a decrease in cerebral blood flow at stenosis greater than the critical as illustrated in Figure 7b, is adequate to maintain normal cerebral blood flow but no reserve blood flow as in Figure 7c, or is adequate to both maintain normal cerebral blood flow and provide reserve even at carotid occlusion as in Figure 7d. These are the model solutions when Pa = 100 mmHg and patient specific collateral vascular resistance values Rw = 3.0, 1.0 and 0.5, the clinically measured mean and +/-SD when carotid stump pressures Ps = 50 +/_ 25mmHg (12).

**Figure 7a,b,c,d.**
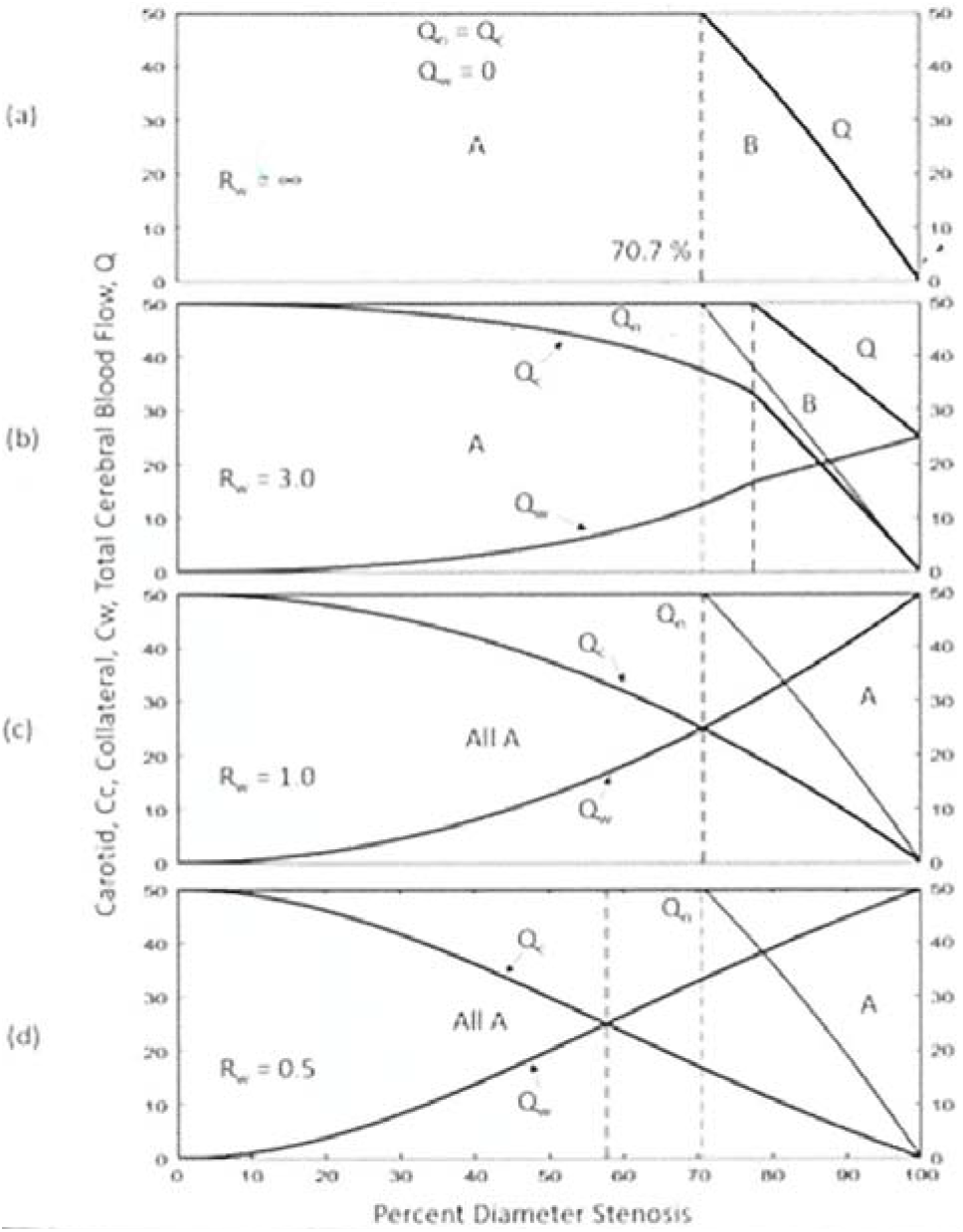
The carotid, Qc, and collateral, Qw, components of cerebral blood flow, Q = (Qc + Qw), as a function of percent diameter stenosis. Systemic arterial pressure is 100 mmHg. Higher or lower systemic arterial pressure values, Pa, will shift the curves slightly. (a). Carotid blood flow in the absence of collateral blood flow. Critical carotid stenosis is 70.7% diameter. (b), (c) and (d). Patients with collateral vascular resistance values Rw = 3.0, 1.0 and 0.5 respectively. These are the mean (Rw = 1.0) plus and minus one standard deviation values. Approximately 16% of patients have a collateral vascular resistance, Rw, greater than 3.0, (b), with poor collateral blood flow and are at risk for significant cerebral ischemia with severe stenosis. Patients with Rw equal to or less than 1.0, (c) and (d), have adequate collateral flow to maintain normal cerebral blood flow when they remain in the autoregulatory zone A. If cerebral perfusion pressure becomes less than 50mmHg Lassen threshold patients with severe stenosis are predicted to transition into ischemic zone B.

The solutions in Figure 7 for carotid blood flow in the autoregulatory zone A and pressure dependent zone B are determined by two model equations. In autoregulatory A zone Q = Qn, where Qn = Qc + Qw and RcQc = RwQw. The solution for carotid blood flow is Qc = Qn/[(Rc/Rw) + 1.0)], where Qn = 50ml/min/100g. In the pressure dependent B zone, Qc = Qcc(1 - X)/(1 - Xc) where Qcc is the carotid blood flow value at the intersection of the two zones and Xc is the fractional percent stenosis at the Qc interface.

Systemic arterial pressure of 100mmHg is used in Figure 7 for simplicity. This Pa value means that the average patient’s collateral vascular resistance is Rw = 1.0 (Figure 7c) and collateral reserve is completely utilized at carotid occlusion. A systemic arterial pressure lower than 100mmHg, like Pa = 87mmHg, the mean in a large series (12), will shift the Rw = 1.0 curve intersection with carotid occlusion (X = 1.0) slightly below the 50 ml/min/100g normal blood. The model predicts that patients with good to excellent collateral vascular resistance, low values Rw 0.5 to 1.0, have a higher proportion of cerebral blood flow from the collateral circulation than from the carotid at moderate to severe carotid stenosis. If this prediction relationship is clinically valid, it explains in part why many patients with carotid occlusion remain asymptomatic.

### Reynolds Numbers

In the 19^th^ century it was recognized that increasing fluid flow rates in experiments led to disturbed flow followed by turbulence negating the accuracy of laminar flow Poiseuille equations. In 1883 Osborne Reynolds reasoned that turbulence occurs when the inertial forces in a fluid become greater than the viscous forces. From this ratio he devised a dimensionless number to predict the transition from laminar to turbulent flow.

The definition of Reynolds number is Rn = VD/kv, where kv is the kinematic viscosity of blood, approximately 3.8 × 10^−2^ (15), V is mean velocity and D is diameter. Assuming an average hemisphere weight of 550g, normal regional cerebral blood flow of 50ml/min/100g becomes 275 cm^3^/min = 4.6 cm^3^/sec. For an average 5mm diameter internal carotid artery with no stenosis Rn = 308. Figure 8 gives the Reynolds numbers for collateral vascular resistance Rw = 1.0, the mean patients value. These results suggest a transition to turbulence between Rn of 400 to 500. Because the model is steady flow these results may be compared to venous flow. One reported value is Rn = 360 in a 3.4mm diameter vein (15).

**Figure 8.**
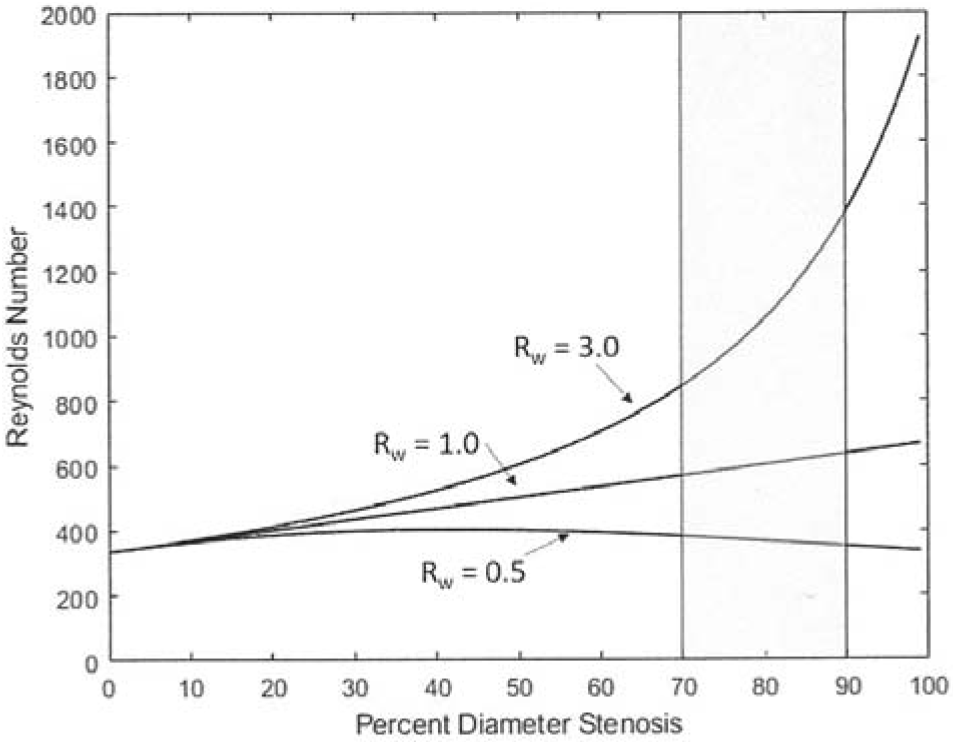
Reynolds numbers for carotid stenosis blood flow when mean arterial pressure is 100 mmHg. The large variance at severe stenosis is proportional to the ratio Qc/Qw. The meaning of Rn numbers for arterial stenosis is not underdtood.

The Lassen model of brain autoregulation negates the ability to obtain a valid Reynolds number for steady flow in a carotid stenosis. However, a turbulence transition Reynolds number can be estimated from the clinical observations of highly disturbed carotid blood flow and cervical bruit in patients at approximately 50% diameter stenosis. Using the estimated hemisphere blood flow of 4.6 cm^3^/sec and a stenosis diameter of 2.5mm in a 5mm diameter internal carotid artery (50% diameter stenosis) gives Rn = 616. This is consistent with theoretical estimates (16) and for carotid stenosis modeling (17) where Re = 600 at turbulence onset.

This is an energy model and the predicted model components energy dissipation must equate energy supplied. A fundamental problem with modeling the effect of carotid artery stenosis on cerebral blood flow is the development of turbulent blood flow at some degree of carotid stenosis. The Poiseuille, Bernoulli, Womersley and other laminar flow solution models fail with the onset of turbulent blood flow (18). The hemodynamics of arterial stenosis is much more complex than blood flow in a non-stenotic artery.

McDonald (15) recognized this and considered a Reynolds number of 300 to 600 to predict the onset of turbulence in an arterial stenosis with pulsatile blood flow.

### Carotid Stenosis Blood Flow Velocity

Figure 7 gives the model predictions of carotid artery stenosis blood flow for a range of normal human vascular resistance values and systemic blood pressures. The law of conservation of mass applied to an incompressible fluid like blood requires the velocity in an arterial stenosis to be average volume blood flow divided by the stenosis lumen area. That is, Vc = Qc/A where Vc and Qc are the mean carotid blood velocities and volume flow and A is the stenosis lumen area. For this study the normal internal carotid diameter, D, is assumed to be 5mm and stenosis lumen area is _π_[D(1 - X)]^2^/4.

Using the carotid volume blood flow values, Qc, in Figure 7, the companion carotid blood flow velocity values, Vc, are given in Figure 9. As with carotid blood flow, velocitie is highly dependent on collateral vascular resistance. The relationship between carotid blood flow velocity between 50% and 80% diameter stenosis is of clinical interest because of the frequent use of Doppler ultrasound velocities to predict degree of stenosis. Above 80% stenosis the velocities are predicted to become highly variable between patients. This is consistent to the findings of Spencer and Reid (18) in their 1979 publication using continuous-wave Doppler ultrasound. They conclude that above 80% diameter stenosis ultrasound frequencies are disrupted by severe turbulence. While severe turbulence may be a major problem, the predicted velocity variance with patient specific collateral vascular resistance between 50% and 80% carotid diameter stenosis indicates that the degree of stenosis predicted by ultrasound velocities may over estimate the degree of carotid stenosis in patients with good collateral vascular resistance (Rw < 1.0) and underestimates it in patients with poor collateral resistance (Rw > 1.0). The turbulent velocity vectors crescendo with increasing stenosis with the resultant forward flow velocity vector being a progressively smaller fraction of the summed velocity vector Reynolds stresses of turbulent flow (16).

**Figure9.**
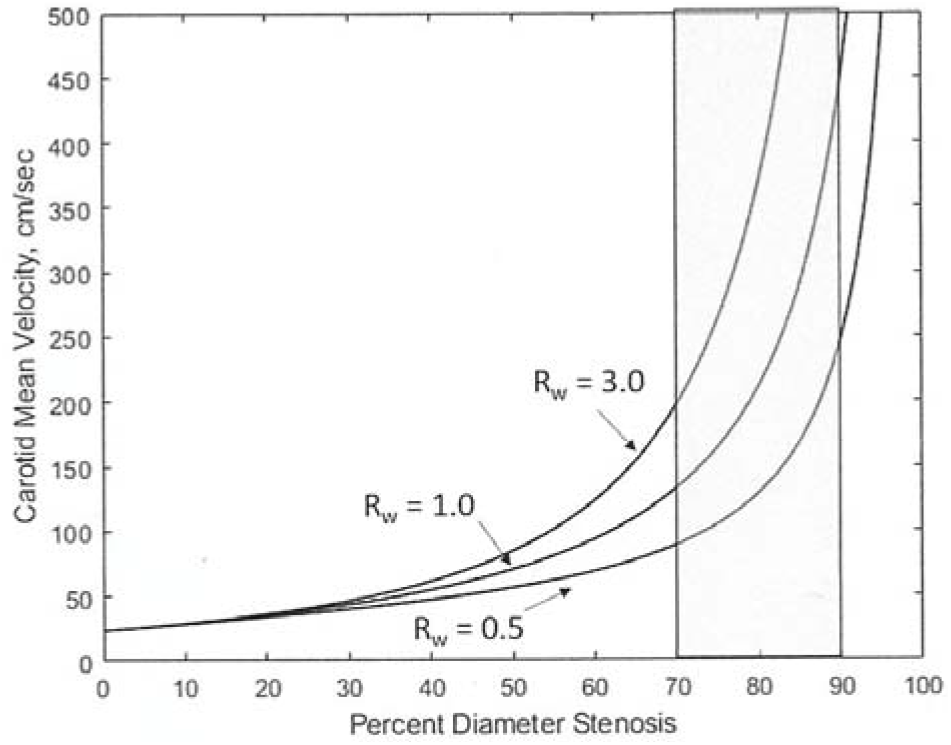
Model predictions for carotid stenosis mean velocities. Patient specific collateral vascular resistance has an increasing impact on velocity with progressive carotid stenosis. These variance in mean velocity results raise concerns regarding the accuracy of ultrasound carotid stenosis velocities and predicted degree of stenosis.

The model carotid blood flow predictions are based on steady, not pulsatile blood flow.

Hemodynamic analysis of pulsatile arterial blood flow is far more complicated than steady flow because the pressure and flow wave forms generated by left ventricular ejection are not in phase (time). The ratio of pulsatile pressure to pulsatile flow is defined as impendence, a complex concept. The internal carotid input volume blood flow profile is characteristic of solid organs with peak flow approximately 30% to 40% greater than mean flow. Applying this simplistic approach, carotid artery blood flow velocities are approximately 1.3 to 1.4 times the values in Figure 9.

### Carotid Stenosis Energy

The total energy per unit time supplied to the cardiovascular system is by definition the systemic pressure times cerebral blood flow. The total model energy E is PaQ, or E = PaQn, when cerebral flood flow is normal. Because Qn is cerebral blood flow per 100grams of brain these values are energy density per minute. This is actually power density but because time is not a variable in steady flow energy is also correct. The law of energy conservation model equations optimize/minimizes energy utilization, as in nature. Normalizing the equations to zero residual energy at brain blood flow exit means that the total input energy, PaQ, is utilized by the model. Each of the three components (carotid, collateral and brain) contributes to optimize and minimize energy consumption. The distribution of energy density between components varies with the degree of carotid stenosis. For example, with no stenosis, Rc = 0, the perfusion pressure, Pp, is systemic pressure, Pa, and all energy is dissipated by the brain. With carotid occlusion the energy utilized by the collateral circulation, Ew = Qw(Pa - Pp), and by the brain, Eb = QwPp, and total energy density is E = Ew + Eb. Total blood flow, Q, is Qw, not necessarily Qn unless collateral flow is adequate to maintain normal cerebral blood flow as in Figure 7 d when Rw is greater than 1.0 at carotid occlusion. With carotid stenosis the distribution of energy dissipation between the carotid and collateral components is complex and a function of Pa, Rw and Rc, as illustrated in Figures 7 b, c and d.

Carotid blood flow is inversely proportional to the availability of collateral blood flow as predicted by Rw values. Figure 10 is the predicted carotid energy utilization as a function of percent diameter stenosis. Similar to the carotid blood flow distributions given in Figure 7, the carotid energy dissipation is significantly higher in patients with very poor collateral circulation, Rw values greater than 3.0.

**Figure 10.**
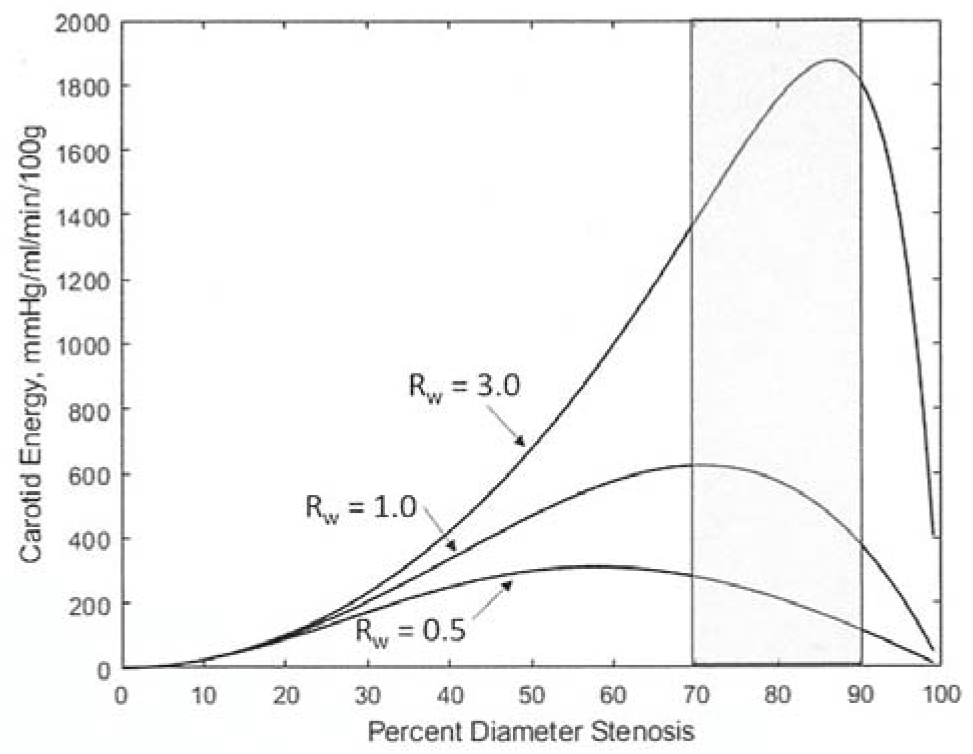
Carotid stenosis energy dissipation at mean arterial pressure of 100 mmHg. Patients with poor collateral flow, Rw > 1.0 are predicted to have greater carotid stenosis energy dissipation than those with good collateral flow. This may have implications for carotid plaque damage and stability.

Similar to Figure 9 for carotid artery velocities, patients with poor collateral circulation have the highest carotid stenosis energy dissipation. This is consistent with high degrees of turbulence, more cervical acoustical energy and tissue deformation energy dissipation as well as higher ultrasound measured blood flow velocities. While increasing systemic arterial pressure is predicted to decreases the risk of cerebral ischemia in patients with significant carotid stenosis Figure 7, it also increases carotid stenosis energy dissipation and adverse carotid hemodynamics.

## Discussion

The objectives of this study were to use established models to determine the role of three patient specific independent variables, systemic arterial pressure, collateral vascular resistance and carotid stenosis to predict cerebral blood flow, cerebral ischemia, blood flow reserve, critical carotid stenosis and carotid stenosis hemodynamics. Both the hemodynamic energy conservation model and Lassen cerebral blood flow autoregulation model and their associated algebraic equations are straightforward and basic. Their blood flow solutions are complex and provide new and interesting clinical predictions for patients with carotid artery stenosis. It is a daunting challenge to model a carotid stenosis cerebrovascular system because of turbulent blood flow. The choice is between a simple steady flow energy model and a modern computational fluid mechanics analysis of the complex partial differential equations of pulsatile flow. This study is the former, a one-dimensional energy conservation blood flow model based on the definition of vascular resistance coupled with Lassen cerebral blood flow autoregulation. While the solutions reveal little regarding the nature of turbulent flow, the results may justify the method.

Similar but highly complex three-dimensional energy models have been used to study focal arterial and cardiac valve turbulence (19) and focal carotid bifurcation turbulent flow (20). To date there are no published comprehensive three-dimensional computational hemodynamic studies of carotid stenosis hemodynamics. One computational analysis uses a pulsatile flow and non-Newtonian viscosity model to predict transition of laminar blood flow to turbulence between 50% and 66% diameter arterial stenosis (21). Above this stenosis level accurate computational solutions are missing because the Navier–Stokes equations break down with intense turbulence. The simple model used herein bypasses this problem. The predicted rapid increase in carotid stenosis energy after 50% diameter stenosis, illustrated in Figure 10, is the turbulent energy dissipation produced by the pressure gradient across the stenosis. This ability of this basic energy model to incorporate turbulent blood flow energy loss allows solutions that may have clinical value for patients with moderate to severe carotid artery stenosis.

### Critical Carotid Stenosis

The combined hemodynamic and Lassen models predict critical stenosis values. The standard definition of critical arterial stenosis is the degree at which further stenosis decreases blood flow. For arteries in general, this has little meaning because for every volume flow rate there is a critical stenosis value. The internal carotid artery is unique because the blood flow it supplies to the cerebral hemisphere is normally autoregulated at approximately 50ml/min/100g. Critical carotid stenosis is reached when cerebral perfusion reaches the 50mmHg Lassen threshold. Values range from 50% to 76% diameter stenosis depending on systemic arterial pressure.

Critical carotid stenosis is the key to predicting (a) the relationship between degree of carotid stenosis and carotid and collateral blood flows in patients with moderate to severe carotid stenosis, (b) the adequacy of cerebral blood flow and (c) carotid artery stenosis hemodynamics. In patients with carotid stenosis greater than the critical value there is no reserve carotid blood flow and collateral blood flow must be adequate to maintain normal cerebral blood flow or prevent cerebral ischemia if blood flow decreases.

### Systemic Arterial Pressure

In addition to critical carotid stenosis, systemic arterial pressure is also a primary determinant of the magnitude of collateral blood flow, reserve cerebral blood flow and carotid hemodynamics. These complex solutions are given as a function of percent diameter carotid stenosis in Figures 3 to 10. For numerical simplicity some figures use a systemic pressure of 100mmHg. Several general conclusions can be made from these results. Patients with moderate to severe carotid stenosis and good collateral circulation as measured by low collateral vascular resistance are predicted to have adequate reserve blood flow and low risk of cerebral ischemia at normal systemic arterial pressures. Patients with high collateral vascular resistance are at increased risk but may be protected from cerebral ischemia by mild hypertension. Conversely, systemic hypotension may promote cerebral ischemia.

### Collateral Vascular Resistance

The primary reason why this model has blood flow solutions is a quantitative definition of a patient’s collateral blood flow potential, their collateral vascular resistance. The definition is a model solution when the carotid is occluded. It requires knowledge of a patient mean systemic arterial pressure and carotid stump pressure. Having made and recorded over 1,000 systemic and stump pressure measurements in patients with significant carotid stenosis, the mean and standard deviation values of collateral vascular resistance in this population were calculated. This provides clinically relevant model solutions.

### Carotid and Collateral Blood Flows

The anatomic human and model parallel carotid and collateral hemodynamic systems determine available regional cerebral blood flow. When carotid stenosis is less than critical, cerebral blood flow is primarily from the carotid artery, although patients with low collateral vascular resistance have a progressively higher collateral blood flow contribution. At carotid stenosis greater than critical, collateral blood flow is the primary determinant of cerebral blood flow. Adequate collateral blood flow is essential to prevent cerebral ischemia at severe carotid stenosis or occlusion and is the sole determinant of reserve cerebral blood flow.

### Carotid Velocity and Energy

These model solutions were obtained using the carotid blood flow solutions and should be viewed accordingly. Both carotid blood flow velocity and carotid energy dissipation are strongly influenced by the collateral circulation. The predicted wide variances in these calculated values at high-moderate and severe carotid stenosis are concerning. If these model results even mildly predict clinical reality, in-depth investigations of carotid stenosis blood flow velocities and energy production-loss are needed as suggested below.

### Clinical Implications

The advantage of higher mean systemic arterial pressure is a recurring model prediction. Increasing systemic pressure (a) shifts percent critical carotid stenosis higher, resulting in improved cerebral hemodynamics, (b) increases carotid stump pressure decreasing selective shunt use and (c) decreases the possibility of cerebral ischemia during carotid surgery without a shunt. For example, in a study of 107 patients having carotid endarterectomy with carotid stump pressures greater than 25mmHg (no shunt), 26 required postoperative nitropresside or nitroglycerin to maintain systolic blood pressure below 160mmHg. The mean carotid stump pressure in this group was 36mmHg compared to 46mmHg in the 81 patients not requiring postoperative hypertension treatment (P<0.01) (22). While the mean systemic arterial pressure was not measured at the time of stump pressure measurement, it is highly likely that the collateral vascular resistance values were significantly higher in the hypertension cohort. This suggest that mild cerebral ischemia during carotid occlusion may trigger a systemic hypertension response. Similarly, mild hypertension may be protective from cerebral ischemia due to significant carotid stenosis as model solutions predict.

The clinical implications for patients with asymptomatic moderate to severe carotid artery stenosis are significant if the model predictions approximate actual cerebrovascular hemodynamics. While normal cerebral blood flow is auto-regulated, the relative contributions from the carotid and collateral blood flow components are highly dependent on their respective vascular resistances. When carotid stenosis is less than critical, cerebral blood flow is normal but the ratio of carotid to collateral blood flow is dependent on collateral vascular resistance. In theory, noninvasive measurements of mild to moderate degree of carotid stenosis (less than critical) and carotid blood flow could be used to estimate patient specific collateral resistance. Unfortunately, these measurements are not accurate enough to do so.

Measurements of cerebral reserve or reactivity in patients with less than critical carotid stenosis are invalid because reserve blood flow has both carotid and collateral components. Conversely, at carotid stenosis greater than a patient’s critical stenosis value, all reserve blood flow is collateral. The model solution for collateral vascular resistance at stenosis less than 100% (occlusion) but greater than critical, requires measurements of three patient specific variables, systemic pressure, percent stenosis and reserve blood flow. This offers an opportunity to determine patient specific collateral vascular resistance without knowing carotid stump pressure. Absent or minimal reserve is a predictor of inadequate collateral blood flow to prevent cerebral ischemia. While measurement of reserve regional volume blood flow is more accurate and precise than cerebral reactivity, the breath-holding trans-cranial Doppler velocity reactivity index may be the simplest way to determine collateral vascular resistance and identify the sub-set of patients with asymptomatic significant carotid stenosis that would benefit from carotid endarterectony.

Both predicted carotid stenosis velocities and energy dissipation have potential clinical implications. Doppler ultrasound velocity measurements are the current non-invasive standard for screening and following patients with carotid stenosis. Carotid velocity values have little clinical meaning prior to the onset of turbulent blood flow near 50% diameter stenosis. At degrees of stenosis above critical the increasing variance of patient specific carotid velocity magnitude raises the question of the accuracy and precision of ultrasound determined degree of carotid stenosis. These velocity values are calculated from the model predictions of carotid blood flow at specific carotid stenosis values. An accurate measurement of carotid stenosis minimum diameter or area is needed to determine the validity of ultrasound derived percent stenosis. Assuming a normal internal carotid artery diameter to be 5mm, a 60% diameter stenosis has a 2mm lumen.

Current anatomic imaging methods have difficulty with diameters at and below 1mm, or 80% diameter stenosis.

The model predictions for carotid stenosis energy dissipation are also clinically concerning. Both blood flow velocity and carotid energy dissipation are strongly influenced by the collateral circulation. It has been suggested that poor collateral circulation resulting in high carotid energy dissipation adversely affects plaque stability (23). Recently the effect of high-grade carotid stenosis on the pressure gradient produced by turbulence has been estimated using 4D Flow MRI to measure energy dissipation (24). Carotid energy dissipation increases in intensity and variability with progressive stenosis in proportion to a patient’s collateral vascular resistance. If focal carotid blood flow energy dissipation plays a significant role in plaque stability, patients identified with high plaque energy dissipation are at increased risk of embolization and may be considered for carotid surgery independent of their current degree of carotid stenosis.

Prior to the dominance of duplex ultrasound for non-invasive carotid evaluation, multiple techniques were used to estimate the significance of carotid stenosis including cervical bruit acoustic measurements. Perhaps this technology was abandoned too soon. The model predicts a wide between patient variance in carotid volume blood flow, blood flow velocity and turbulent energy dissipation. This is crescendo when carotid stenosis is greater than critical. Turbulent energy dissipates from blood into the carotid walls and surrounding tissue with mechanical deformation/strain, vibration/oscillation at various frequencies and heat. Some components of this complex process can be measured acoustically and possibly thermally to estimate the influence of collateral vascular resistance on a patient’s risk of hemodynamic or embolic stroke.

### Limitations

The two major limitations of this study are the use of linear or one-dimensional models to predict blood flows and the assumption of steady blood flow. Reality is pulsatile blood flow with out-of-phase pressure and velocity/volume waveforms. The model results are cardiac cycle time average values. Applying a pressure or velocity/volume waveform to model mean flow results may have utility but difficult to justify. On balance, pulsatile cerebral and collateral blood flow is damped being constrained in a rigid skull. With the exception of cervical carotid stenosis mean blood flow model solutions may be reliable.

The standard hemodynamic energy conservation model for vascular systems as applied to the cerebral circulation includes two additional models. The Lassen cerebral autoregulation model is used to define cerebral vascular resistance and the threshold for reduced cerebral blood flow. Carotid vascular resistance is assumed to be proportional to fractional percent stenosis lumen area. This is identical to carotid vascular resistance being proportional to fractional percent stenosis diameter squared. The third model component, collateral vascular resistance, is determined by the model solution when the carotid is occluded and cerebral perfusion pressure equals carotid stump pressure, a clinical measurement. Models do not define nature but rather predict functional relationships including the roles of patient specific independent variables. In this study cerebral blood flow and its carotid and collateral components are dependent variables.

The study aims to predict blood flow as a function of carotid stenosis, which is both an independent variable and patient specific. The other two key independent and patient specific variables are systemic arterial blood pressure and collateral vascular resistance. While each of the three model components is anatomically distinct in humans.

The second major assumption is steady, not pulsatile, blood flow. As stated above, pulsatile pressure and flow waveforms are not generally in phase. This means that simple steady flow resistance becomes complex impedance with pulsatile flow. The blood flow results in this study are the time average over a standard cardiac cycle of blood flow and pressure and results should be interpreted with this in mind. On balance, cerebral blood flow occurs inside the skull, a rigid structure that dampens flow waveforms, similar to that observed in solid organs.

Other potential limitations include assuming a normal value for regional cerebral blood flow and a lower threshold cerebral perfusion pressure for auto-regulation. Assumed cerebral arterial pressures are in the normal range. The calculated mean and standard deviations for collateral vascular resistance in patients with significant carotid stenosis are based on clinical measurements. Using different assumed values will change the numerical values of the examples given in the figures but not the general trend or concept.

### Conclusions

Critical carotid stenosis values are defined and predicted by the model. When carotid stenosis is less than critical carotid blood flow is the primary contributor to cerebral circulation. Conversely at higher degrees of stenosis collateral circulation is the main determinant of cerebral blood flow and the sole contributor to reserve blood flow. Both the magnitude of systemic arterial pressure and collateral blood flow as defined by collateral vascular resistance are major determinants of carotid artery blood flow and associated carotid stenosis hemodynamics as a function of percent diameter stenosis.

These are model solutions, not in-vitro measurements, and clinical implications should be considered accordingly.

## Data Availability

all data is published and referenced

## Acknowledgments

Sarah Ellen Archie for manuscript preparation and support and Sujal Dave for figures.

